# Comparison of Very Short Answer Questions and Multiple Choice Questions in Medical Students: Reliability, Discrimination, Acceptability and Effect on Knowledge Retention

**DOI:** 10.1101/2022.07.13.22277583

**Authors:** Roemer J. Janse, Elise V. van Wijk, Bastian N. Ruijter, Jos H.T. Rohling, Jolein van der Kraan, Stijn Crobach, Mario de Jonge, Arnout Jan de Beaufort, Friedo W. Dekker, Alexandra M.J. Langers

## Abstract

**Introduction:** Multiple choice questions (MCQs) offer high reliability and easy machine-marking, but allow for cueing and stimulate recognition-based learning. Very short answer questions (VSAQs) may circumvent these limitations. We investigated VSAQ reliability, discriminative capability, acceptability, and knowledge retention compared to MCQs.

**Methods:** Dutch undergraduate medical students (n=375) were randomised to a formative exam with VSAQs first and MCQs second or vice versa in two courses, to determine reliability and discrimination. Next, acceptability (i.e., VSAQ review time) was determined in the summative exam. Knowledge retention at 2 and 5 months was determined by comparing score increase on the three-monthly progress test (PT) between students tested with VSAQs and students from previous years tested without VSAQs.

**Results:** Reliability (Cronbach’s α) was 0.74 for VSAQs and 0.57 for MCQs in one course. In the other course, Cronbach’s α was 0.87 for VSAQs and 0.83 for MCQs. Discrimination (R_ir_) was 0.27 vs. 0.17 and 0.43 vs. 0.39 for VSAQs vs. MCQs, respectively. Reviewing time of one VSAQ for the entire student cohort was ±2 minutes on average. No clear effect on knowledge retention after 2 and 5 months was observed.

**Discussion:** We found increased reliability and discrimination of VSAQs compared to MCQs. Reviewing time of VSAQs was acceptable. The association with knowledge retention was unclear in our study. This study supports and extends positive results of previous studies on VSAQs regarding reliability, discriminative capability, and acceptability in Dutch undergraduate medical students.

## Introduction

Assessment in medical education commonly uses multiple choice questions (MCQs). Even though this question type offers high reliability and easy machine-marking, it allows for cueing (i.e., answering questions based on cues in the question or answer options rather than on content knowledge) [1-4] and stimulates a recognition-based study approach [5-8]. Although recognition may be sufficient to pass a MCQ-based assessment, it has been critically noted that clinical practice does not offer a multiple choice list of possible diagnoses or procedures, nor is there a single best recognisable answer in the medical profession [9, 10].

Although other question formats have been proposed to circumvent the limitations of MCQs, such as uncued questions and extended matching questions [10-12], these question formats may still facilitate a recognition-based study approach. However, very short answer questions (VSAQs), a free-response type of questions with the answer being limited to 1-4 words, may be better suited to circumvent some of the general limitations of MCQs. The open-ended nature of the VSAQ reflects the clinical practice better and may prevent surface-level study approaches and cueing [1, 3, 4, 8]. In addition, VSAQs are better able to discriminate between students based on proficiency in the content knowledge [1-4, 13, 14] and may increase retention of knowledge [15-17].

Nonetheless, as studies on VSAQs have mainly been performed in the UK, there is a need for validation of previous results in different educational systems [2, 3, 8, 14, 18]. Therefore, we aimed to investigate the reliability, discrimination, acceptability, and retention of knowledge of VSAQs compared to MCQs in a cohort of Dutch medical undergraduate students, partially replicating the design of Sam *et al*. [2]. In addition, we aimed to explore the impact of VSAQs on cueing effects, perceived alignment between assessment and teaching, and student experiences of assessment.

## Methods

### Setting

This study was performed among students partaking in the first year fundamental course “*Regulation and Metabolism*” (RM, May 2021) or in the second year clinical course *“Diseases of the Abdomen”* (DA, April 2021) in the bachelor of Medicine at the Leiden University Medical Center (LUMC), the Netherlands. These courses (6 and 7 weeks, respectively) cover metabolic and gastrointestinal topics. Near the end of the course, students are offered a formative exam and the course ends with a summative exam. After the summative exam, students can evaluate the course with the Automated Education Evaluation System (AEES, **Figure 1A**), which includes questions on constructive alignment. Relevant AEES questions for this study were answered using a 5-point Likert-scale (strongly disagree, disagree, neutral, agree, strongly agree). The LUMC uses RemindoToets (Paragin) [19] for digital assessment with the possibility of proctoring.

**Figure 1.**
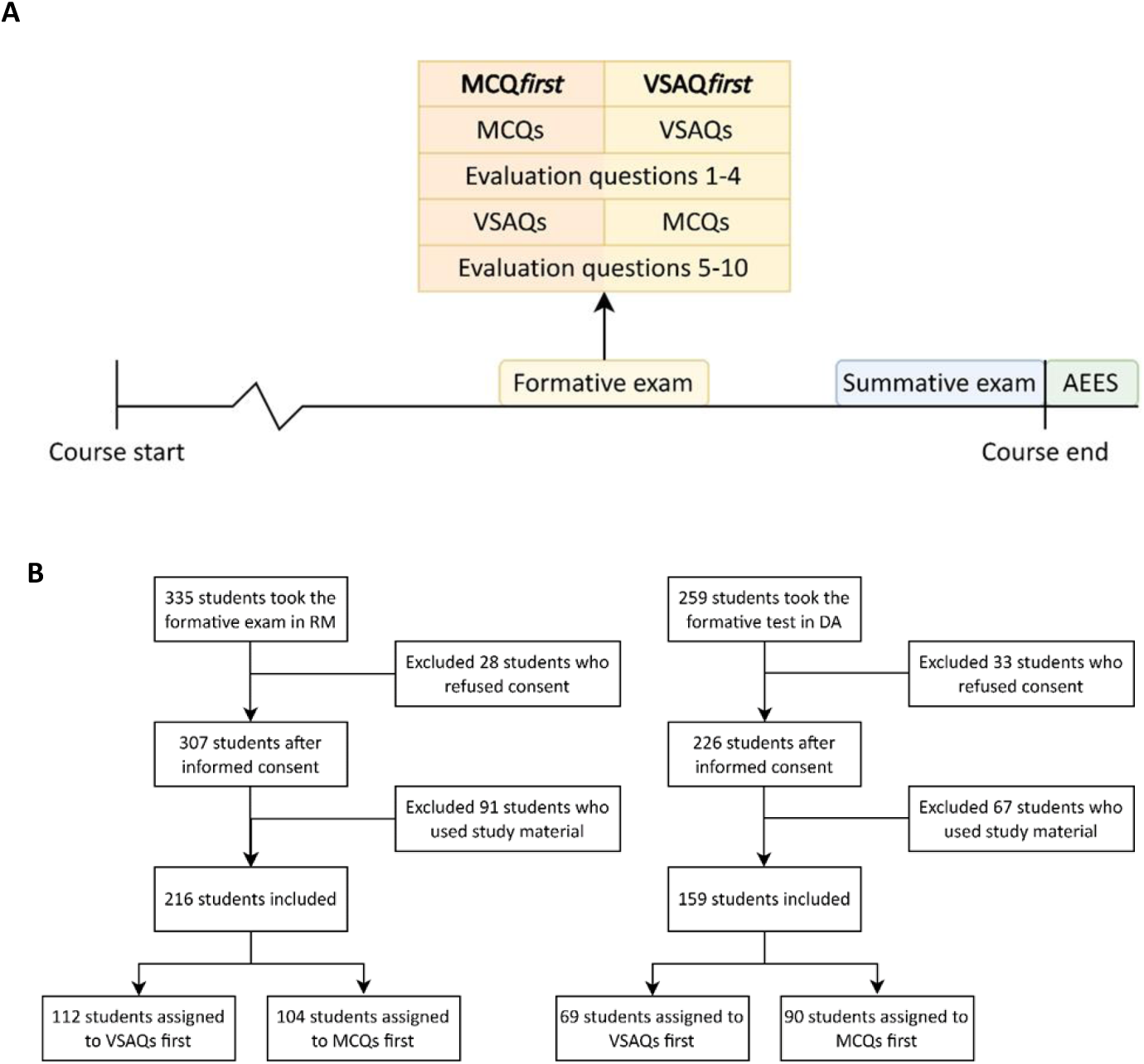
**(A)** Set-up of both courses (RM and DA) with the formative exam and contents, summative exam and contents, and the Automated Education Evaluation System (AEES) **(B)** Flowchart of the study participants.

Next to course-specific examinations, all medical students in the LUMC also participate in a national three-monthly progress test (PT) [20]. The PT consists of 200 MCQs, covering a wide variety of medical topics. Content of the test is identical for all students. Pass and fail thresholds are based on a national benchmark of the results per study-year.

### Formative exam

To determine reliability, discrimination, cueing effects, and students’ insights in the formative exam, students were randomly assigned to a group starting with either MCQs (RM-MCQ*first* and DA-MCQ*first*) or VSAQs (RM-VSAQ*first* and DA-VSAQ*first*), followed by identical questions in the opposing format, similar to the study of Sam *et al*. [2] (**Figure 1A**). The formative exam was available in a fixed timeslot. Participation in this formative exam was mandatory in RM and optional in DA. Exam format order per student was determined using a random number generator in Microsoft Excel (i.e., a Mersenne Twister algorithm) [21]. For DA, 24 completely identical questions in both formats were asked. For RM, 25 questions that tested the same knowledge were asked, albeit sometimes worded differently. Only students who gave informed consent were included in the analysis.

After having finished the first part of the formative exam (either MCQs or VSAQs) students were asked to rate four statements on a 5-point Likert scale (strongly disagree, disagree, neutral, agree, strongly agree) based on the questions they just finished: 1) *The questions are a good representation of how I would be expected to answer questions in clinical practice;* 2) *I found the questions easy;* 3) *I was often unsure whether my answer would be correct)*; 4) *If I had to give an estimate of the grade I would have achieved based on these questions, my estimate would be <grade>*. At the end of the second part of the formative exam, students were asked to rate six more statements: 5) *VSAQs are easier than MCQs*; 6) *VSAQs are more in line with daily clinical practice than MCQs;* 7) *I prepare differently for an assessment with VSAQs than for an assessment with MCQs;* 8) *VSAQs would be a better preparation for clinical practice than MCQs*; 9) *Through the use of VSAQs, the test is better aligned with this course, than a test using MCQs;* and 10) *Any comments I would like to add: <open question>*. Finally, students were asked whether they used study material during the formative exam. Students who did were excluded from the analysis.

Reliability was determined using Cronbach’s α [22], discriminative capability for content knowledge of the items using the R_ir_-value [23], and the average score, calculated over MCQs and VSAQs separately, was stratified by whether students took MCQs or VSAQs first. Cueing effects were determined by comparing whether a student answered the MCQ correct and the equivalent VSAQ incorrect (positive cueing) or whether a student answered the MCQ incorrect and the equivalent VSAQ correct (negative cueing). Although it might have been of influence, the probability of guessing the right answer could not be taken into account. Students’ insights were determined from the evaluation questions asked midway through and at the end of the formative exam.

### Summative exam

The summative exams of RM and DA were rewritten to replace part of the MCQs with VSAQs (45 in RM and 16 in DA). For RM, this was done through rewriting existing MCQs, whereas for DA, a 2-hour workshop was organised for teachers on how to write VSAQs. At the end of both exams, preapproved answers to the VSAQs were automatically marked as correct. Subsequently, teachers reviewed all incorrect answers and could easily add answers that were not in the predefined list, but were also found to be correct. VSAQ review time per question was recorded in DA to determine acceptability. Due to incomplete data collection regarding reviewing time during the initial data collection, these data were collected again one year later. The AEES questionnaire was supplemented with two questions regarding students’ insights: 1) *Because I knew that I would be tested by very short answer questions, I studied in another way than I normally would*; and 2) *Through the use of very short answer questions, the test was a better representation of what I learned in this course, compared to a test using multiple-choice questions*. In addition, the perceived alignment between teaching and assessment was compared to the alignment of the course in the years 2017, 2018, and 2019, determined from two pre-existent questions in the AEES questionnaire: 1) *The assessment as a whole (form and content) is appropriate for what you should have mastered at the end of the course*; and 2) *The (online) test formats (e*.*g. MCQs, open questions, oral and written presentations, practical assessments) matched what I have learned*. Due to emergency remote teaching during COVID-19, 2020 is not considered in these comparisons.

### Progress test

To determine knowledge retention, we studied percent-point score increases on gastrointestinal-related questions in the PT. For 2 months retention, we included the two PTs before and the PT after the course with VSAQs in the analysis. For 5 months retention, we also included the second PT after the course. We compared percent-point score increases on the PT between students from DA who were tested with VSAQs in their summative exam and students from DA who took the course in 2017, 2018, and 2019 (therefore not tested with VSAQs). In a sensitivity analysis, we studied retention in RM. Students from RM were excluded from the main analysis as educational methods of the course had changed considerably compared to the years prior.

### Statistical analysis

Continuous variables are presented as mean (standard deviation) or median (interquartile range) depending on their distribution. Categorical variables are presented as number (proportion). Knowledge retention was determined using a linear mixed model using the restricted maximum likelihood method. The model is adjusted for age, sex, average score increase over previous progress tests, and average bachelor grade on the date of the PT. All statistical analyses were performed using R version 4.1.0 (R Foundation for Statistical Computing, Vienna, Austria).

### Ethical approval

This study was reviewed and approved by the Educational Research Review Board of the Leiden University Medical Center (file number: OEC/ERRB/20201208/1).

## Results

Of the 335 students who took the formative exam in RM, 216 students were included in our study. In DA, 159 of the 259 students who took the formative exam were included (**Figure 1B**). In RM, 104 students started with MCQs (RM-MCQ*first*) and 112 students started with VSAQs (RM-VSAQ*first)*. In DA, 90 students were assigned to DA-MCQ*first* and 69 students to DA-VSAQ*first*.

### Reliability and discrimination

We compared the VSAQs of students starting with VSAQs with the MCQs of students starting with MCQs. This comparison reflects the results of the VSAQs and MCQs that are not influenced by prior questions. VSAQs had higher reliability compared to MCQs (Cronbach’s α 0.74 vs. 0.57 in RM; 0.87 vs. 0.83 in DA for VSAQs vs. MCQs, respectively) (**Table 1**). In the same students, discrimination (mean [SD]), expressed as the R_ir_-value, was higher in VSAQs compared to MCQs (0.27 [0.15] vs. 0.17 [0.13] in RM; 0.43 [0.10] vs. 0.39 [0.10] in DA, for VSAQs vs. MCQs, respectively). The mean scores (mean [SD]) were lower and had a wider distribution width for VSAQs compared to MCQs (57.0 [15.7] vs. 71.2 [12.2] in RM; 51.6 [23.9] vs. 70.0 [19.7] in DA, for VSAQs vs. MCQs, respectively). These results were similar when comparing results within groups (e.g., VSAQs vs. MCQs within MCQ*first*).

**Table 1.** Cronbach’s alpha, average R_ir_ score and mean (SD) scores for the MCQs and VSAQs in MCQ*first* and VSAQ*first*.

|  | Regulation and Metabolism |  |  |  | Diseases of the Abdomen |  |  |  |
| --- | --- | --- | --- | --- | --- | --- | --- | --- |
|  | MCQfirst |  | VSAQfirst |  | MCQfirst |  | VSAQfirst |  |
|  | MCQ<br>(n = 104) | VSAQ<br>(n = 104) | VSAQ<br>(n = 112) | MCQ<br>(n = 112) | MCQ<br>(n = 90) | VSAQ<br>(n = 85) | VSAQ<br>(n = 69) | MCQ<br>(n = 64) |
| Cronbach's $\alpha$ | 0.57 | 0.61 | 0.74 | 0.71 | 0.83 | 0.90 | 0.87 | 0.71 |
| Average $R_{ir}$ | 0.17 | 0.19 | 0.27 | 0.27 | 0.39 | 0.49 | 0.43 | 0.26 |
| (SD) | (0.13) | (0.14) | (0.15) | (0.09) | (0.10) | (0.13) | (0.10) | (0.10) |
| Mean score | 71.2 | 72.3 | 57.0 | 75.4 | 70.0 | 58.4 | 51.6 | 72.5 |
| (SD), % | (12.2) | (12.9) | (15.7) | (14.1) | (19.7) | (25.9) | (23.9) | (15.0) |

### Acceptability

In the initially collected data, the average reviewing time per VSAQ by one teacher in the summative exam of DA (7 VSAQs, 308 students) was 2 minutes and 20 seconds (SD 52 seconds). Additionally, on average 2 minutes and 9 seconds (SD 2 minutes and 36 seconds) were spent replying to comments and consultation of other teachers. The maximum time spent on a single VSAQ was 11 minutes and 24 seconds. Because of incomplete data collection, data were collected again during the next course of DA. One year later (22 VSAQs, 338 students), the average time spent on reviewing questions in DA was 1 minute and 58 seconds (SD 40 seconds) and consultation of other teachers took on average 36 seconds (SD 47 seconds).

### Retention

The linear mixed model used to determine the knowledge retention in students from DA yielded an average percent-point score increase, adjusted for baseline score increase (PTs 1-5), age, sex, and average bachelor grade, of -1.1 (95% CI -3.3, 1.1) per test for 2 months retention among students assessed with VSAQs compared to students from previous years who had been assessed with MCQs (**Supplemental table 1**). For 5 months retention, this was -2.5 (95% CI -4.2, -0.8) per test. The sensitivity analysis in which we fit the model in RM showed an increase of 3.1 (95% CI 2.0, 4.3) for 2 months and 3.8 (95% CI 2.8, 4.8) for 5 months retention.

### Secondary outcomes

Positive cueing, defined as a correctly answered MCQ with an incorrectly answered equivalent VSAQ, occurred on average more often per student in RM-VSAQ*first* and DA-VSAQ*first* (20.0%, IQR; 16.0-28.0%-29.2%; 20.8%, IQR; 12.5-29.2%, respectively) compared to RM-MCQ*first* and DA-MCQ*first* (4.0%, IQR; 4.0-8.0%; 8.3%, IQR; 4.2-16.7%, respectively) (**Supplemental table 2**). On a question level, positive cueing occurred in 100% of questions in all groups. The frequency of positive cueing per question was on average higher in RM-VSAQ*first* and DA-VSAQ*first* (14.3%, IQR 7.1-33.9%; 22.7%, IQR 10.9-28.5%, respectively) compared to RM-MCQ*first* and DA-MCQ*first* (4.8%, IQR 2.9-9.6%; 15.9%, IQR 11.8-20.3%, respectively) (**Supplemental table 3**). Negative cueing in students, which was defined as students answering the VSAQ correctly and the equivalent MCQ incorrectly, occurred more often per student in RM-MCQ*first* compared to RM-VSAQ*first* (8.0%, IQR 4.0-12.0% to 4.0%, IQR 0.0-4.0%). In DA-MCQ*first*, negative cueing was on average not observed in students (0.0%, IQR 0.0-4.2%). Negative cueing per question occurred in 92%, 56%, 79%, and 79% of the questions for RM-MCQ*first*, RM-VSAQ*first*, DA-MCQ*first*, and DA-VSAQ*first*, respectively. The frequency of negative cueing per question was on average lower in RM-VSAQ*first* compared to RM-MCQ*first* (0.9%, IQR 0.0-1.8%; 3.8%, IQR 1.9-12.5%), but higher in DA-VSAQ*first* compared to DA-MCQ*first* (3.1%, IQR 1.6-5.1%; 1.8%, IQR 1.2-3.5%). The maximum percentage of positive cueing by students in a single question was the highest in RM-MCQ*first* (62.5%). The maximum percentage of negative cueing by students in a single question was 38.5% in RM-MCQ*first*.

More than 80% of students were uncertain about answering VSAQs correctly (86% and 82% in RM and DA, respectively) (**Supplemental table 4, Figure 2**). Approximately 90% of students (strongly) disagreed that VSAQs were easier than MCQs (**Supplemental table 5**,**6**). 51% of students in RM and 46% in DA (strongly) agreed that assessment with VSAQs changed their test preparation. In DA, 60% of students agreed or strongly agreed that VSAQs better represented clinical practice. This was 34% in RM. Almost 70% of students in RM and 48% in DA (strongly) disagreed that the test was better aligned with the course by using VSAQs. 45% of students in RM and 42% in DA (strongly) agreed they would change learning behaviour if tested with VSAQs (**Supplemental table 7**). 83% of students in RM (strongly) disagreed that VSAQs better represented what they learned during the course than MCQs. This was 51% in DA. Perceived alignment of assessment, teaching and learning activities are reported in **Supplemental table 8**.

**Figure 2.**
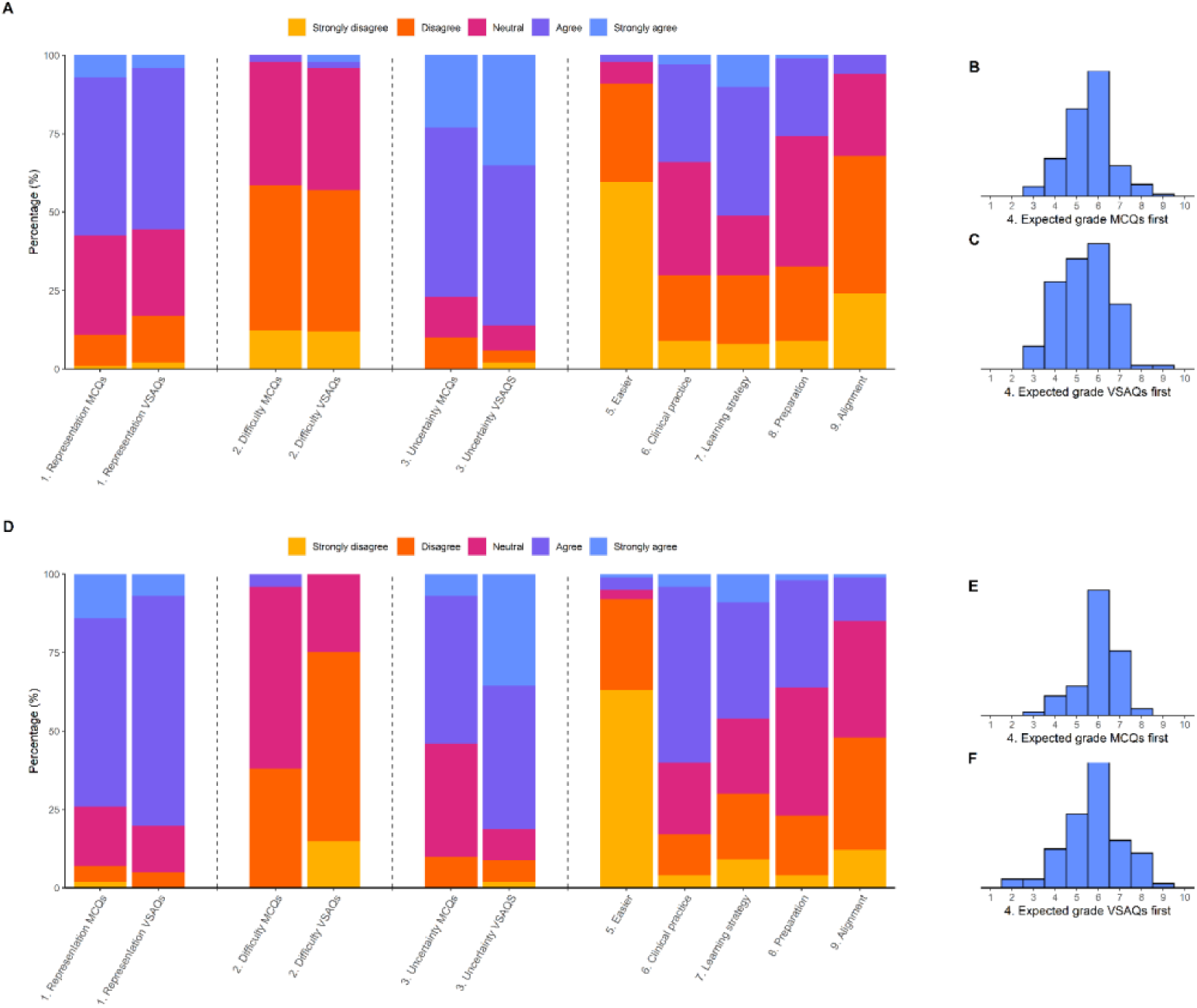
Students’ experiences and grade estimates of the MCQs and VSAQs in the formative exam. Distribution of the answers given to the 5-point Likert scale evaluation questions halfway through the exam after the MCQs or VSAQs and at the end of the exam; and estimates of their grade halfway through the exam in RM **(A, B, C)** and DA **(D, E, F)**.

## Discussion

This study supports and extends earlier work by Sam *et al*. [2] by analyzing VSAQs in a Dutch cohort of medical bachelor students. Results showed higher reliability and discrimination of VSAQs compared to MCQs. Furthermore, in our opinion, the time needed to mark VSAQs was acceptable. In our study, effects on 2 and 5 months retention were unclear. Overall, VSAQs scored better on reliability, discrimination, and perceived alignment in DA but only slightly better in RM, which may be attributable to the workshop offered to the teachers, course material being better suited for VSAQs and the opportunity for students to practice with the VSAQs prior to the exams in DA.

We found higher reliability and discrimination of VSAQs compared to MCQs. Moreover, students scored lower on VSAQs. These findings may in part reflect the decreased possibility of guessing correctly in VSAQs, and are line with Sam *et al*. [2] and other previous studies in undergraduate medical students in the UK [14, 18]. The lower score also suggests that VSAQs are more difficult, possibly due to a need of answer generation, rather than answer recognition. The attenuation of differences in RM-MCQ*first* and DA-MCQ*first* might be explained by cueing and guessing. As VSAQs can largely circumvent cueing and guessing, they provide a better measure of a students’ true content knowledge, which increases validity [2, 8, 14]. The high discriminative capability of VSAQs is further supported by higher average R_ir_ values of VSAQs in DA. In RM, average R_ir_ values were low for both MCQs and VSAQs, which indicates low question performance regardless of format. This difference might be because VSAQs were rewritten from MCQs in RM, but written from scratch in DA.

We considered the reviewing time of VSAQs by teachers using an assessment software program acceptable. This is supported by previous studies that found comparable and shorter review times, using different marking systems, multiple examiners, and more questions [2, 14]. Although our results are derived from the review process of only one examiner and a limited number of VSAQs, it further supports the efficient marking process of VSAQs. Nonetheless, whereas not every MCQ has to be reviewed, it should be noted that a VSAQ should always be reviewed after machine marking, although repeated use of questions may decrease reviewing time, depending on software used [2].

Knowledge retention was compared in DA between students in our study tested with VSAQs and students from previous years not tested with VSAQs. We observed a similar score increase at 2 months, but a lower score increase at 5 months. Nonetheless, in RM, we observed higher average score increases for 2 and 5 months retention. The decrease observed in DA, contrary to the results of RM, may be due to questions in the PT covering more topics discussed in RM compared to DA. Furthermore, transfer-appropriate-processing might apply here, which states that memory is enhanced when the initial encoding and later retrieval of the information are similar [24], because the information is encoded using VSAQs but retrieved in the PT using MCQs. However, other studies have shown information retention occurring regardless of the final test format [15, 25]. Because students had not been sufficiently assessed with VSAQs before and therefore were not used to this question type, they might not have adjusted their learning strategy yet, even though a substantial number of students stated that they would change the way they studied if they were to be assessed with VSAQs. Last, we cannot rule out COVID-19 related online teaching interference with the motivation and education of students [26-28]. Although our study remains inconclusive regarding knowledge retention, other studies have supported the positive effect of VSAQs on long-term retention, indicating higher retention in students taking an initial VSAQ test compared to a MCQ test [15-17], which is most likely explained by the greater level of retrieval difficulty demanded by VSAQs leading to better retention of information [25, 29].

Positive cueing per student occurred more often in the students who started with VSAQs, which is in line with the findings of Sam *et al*. [2]. This is expected, as students answering the VSAQs first and MCQs second cannot carry over the MCQ answer to the VSAQ, therefore having to rely on content knowledge for the VSAQ. Cueing per question was also seen more often in this group, but not for every question [2]. Although positive cueing entails students choosing the right answer based on the available answer options and not content knowledge, we most likely also measured students guessing the right answer, as it is nearly impossible to separate these two effects in MCQs [30]. We observed more negative cueing in RM in MCQ*first* compared to VSAQ*first*, whereas in DA we observed no negative cueing in MCQ*first* compared to 4.3% in VSAQ*first*, which might be related to course content. In Sam *et al*. negative cueing was the same among both groups [2]. In many questions cueing occurred, but per question it was observed only a few students.

We aimed to evaluate the perceived constructive alignment between teaching and learning activities and assessment by comparing the answers to questions of the current and previous years on this topic in a recurrent evaluation questionnaire, the AEES. Students of RM perceived alignment between assessment and teaching lower after being assessed with VSAQs. However, clear interpretation of these data is difficult due to large variation in response rate and significant disruptions of COVID-19 on the education system and students’ learning, as well as adaptations in the course. Students’ experiences in our study are comparable with those found by Sam *et al*. [2]. The vast majority of the students thought the VSAQs were more difficult than MCQs and almost half of the students said they changed their learning behaviour because they were assessed with VSAQs. Noteworthy are differences in experiences between courses. Concerning connection of VSAQs with clinical practice, students of DA were more positive than students of RM, which might be due to differences in course content and quality of the VSAQs. Feedback provided by students mainly concerned VSAQ construction. Moreover, feedback suggested that VSAQ phrasing might not always have been clear enough. This led to uncertainty regarding the level of specificity of the desired answer and highlights the importance of a well-designed VSAQ in order to profit from their advantages over MCQs [8, 31].

Study strengths are the randomised design, studying two different courses and the investigation of both student and teacher perspectives. Limitations are the inclusion of students from only one institute, the relatively small sample size, and interference of the COVID-19 pandemic. Home-based studying can lead to social detachment from fellow students and deterioration in study performance in medical students [32]. This might have negatively affected the effort and motivation of students in their learning process and the assessments. Furthermore, as the formative exam had no consequences for a pass/fail decision of the course, we cannot be certain that students performed at their best when answering the questions. To determine acceptability, we used only one reviewer who logged the times by hand, leading to less accurate reviewing times. To obtain a more precise measure of acceptability, these findings could be extended by using multiple examiners, more VSAQs and automatically logged times.

In conclusion, this study validates the positive results of Sam *et al*. [2] on VSAQs in terms of reliability, discrimination, and acceptability in formative assessments in a Dutch cohort of undergraduate medical students. Wider implementation of VSAQs in medical education seems justified and may improve assessment in medical education.

## Data Availability

All data in the present study are available upon reasonable request to the corresponding author.

