## Supplemental Material for "Comparison of Very Short Answer Questions and Multiple Choice Questions in Medical Students: Reliability, Discrimination, Acceptability and Effect on Knowledge Retention"

*Supplemental materials*

Roemer J. Janse*, Elise V. van Wijk*, Bastian N. Ruijter, Jos H.T. Rohling, Jolein van der Kraan, Stijn Crobach, Mario de Jonge, Arnout Jan de Beaufort, Friedo W. Dekker, Alexandra M.J. Langers.

* These authors contributed equally to this work.

### Supplemental tables

**Supplemental table 1.** Retention of knowledge of students tested with VSAQs compared to students of historical cohorts tested with MCQs. Numbers represent average percent-point score increase per measure moment among VSAQ group compared to MCQ group.

| | **Diseases of the Abdomen** | **Regulation and Metabolism** |
|---|---|---|
| 2 month retention | -1.1 (-3.3, 1.1) | 3.1 (2.0, 4.3) |
| 5 month retention | -2.5 (-4.2, -0.8) | 3.8 (2.8, 4.8) |

**Supplemental table 2**. Positive and negative cueing per person in MCQ*first* and VSAQ*first.*

| | **Regulation and Metabolism** | | **Diseases of the Abdomen** | |
|---|---|---|---|---|
| | **MCQ*first* (*n* = 104)** | **VSAQ*first* (*n* = 112)** | **MCQ*first* (*n* = 90)** | **VSAQ*first* (*n* = 69)** |
| Positive cueing, median (IQR), % | 4.0 (4.0 - 8.0) | 20.0 (16.0 - 28.0) | 8.3 (4.2 - 16.7) | 20.8 (12.5 - 29.2) |
| Negative cueing, median (IQR), % | 8.0 (4.0 - 12.0) | 4.0 (0.0 - 4.0) | 0.0 (0.0 - 4.2) | 4.2 (0.0 - 4.2) |

**Supplemental table 3**. Positive and negative cueing per question in MCQ*first* and VSAQ*first.*

| | **Regulation & Metabolism** | | **Diseases of the Abdomen** | |
|---|---|---|---|---|
| | **MCQ*first* (*n* = 104)** | **VSAQ*first* (*n* = 112)** | **MCQ*first* (*n* = 90)** | **VSAQ*first* (*n* = 69)** |
| **Frequency of questions where cueing occurred, %** | | | | |
| Positive cueing | 100 | 100 | 100 | 100 |
| Negative cueing | 92 | 56 | 79 | 79 |
| **Average frequency of cueing per question, %** | | | | |
| Positive cueing, median (IQR) | 4.8 (2.9 - 9.6) | 14.3 (7.1 - 33.9) | 15.9 (11.8 - 20.3) | 22.7 (10.9 - 28.5) |
| Positive cueing, max | 26.9 | 62.5 | 32.9 | 43.8 |
| Negative cueing, median (IQR) | 3.8 (1.9 - 12.5) | 0.9 (0.0 - 1.8) | 1.8 (1.2 - 3.5) | 3.1 (1.6 - 5.1) |
| Negative cueing, max | 38.5 | 16.1 | 7.1 | 10.9 |

**Supplemental table 4.** Distribution of the answers given to the 5-point Likert scale evaluation questions halfway of the formative exam after MCQs (MCQ*first*) or VSAQs (VSAQ*first)*.

| | **Regulation and Metabolism** | | | | | | **Diseases of the Abdomen** | | | | | |
|---|---|---|---|---|---|---|---|---|---|---|---|---|
| | **MCQ*first* (*n* = 104)** | | | **VSAQ*first* (*n* = 112)** | | | **MCQ*first* (*n* = 85)** | | | **VSAQ*first* (*n* = 64)** | | |
| | EQ1 | EQ2 | EQ3 | EQ1 | EQ2 | EQ3 | EQ1 | EQ2 | EQ3 | EQ1 | EQ2 | EQ3 |
| Strongly disagree | 1% | 12% | 0% | 2% | 12% | 2% | 2% | 0% | 0% | 0% | 15% | 2% |
| Disagree | 10% | 46% | 10% | 15% | 45% | 4% | 5% | 38% | 10% | 5% | 61% | 7% |
| Neutral | 32% | 39% | 13% | 28% | 39% | 8% | 19% | 58% | 36% | 15% | 25% | 10% |
| Agree | 51% | 2% | 54% | 52% | 2% | 51% | 60% | 4% | 47% | 74% | 0% | 46% |
| Strongly agree | 7% | 0% | 23% | 4% | 2% | 35% | 14% | 0% | 7% | 7% | 0% | 36% |

EQ1: *The questions are a good representation of how I would be expected to answer questions in clinical practice.*
EQ2: *I found the questions easy.*
EQ3: *I was often unsure whether my answer would be correct.*

**Supplemental table 5.** Distribution of the answers given to the 5-point Likert scale evaluation questions at the end of the formative exam.

| | **Regulation and Metabolism (*n* = 216)** | | | | | **Diseases of the Abdomen (*n* = 146)** | | | | |
|---|---|---|---|---|---|---|---|---|---|---|
| | EQ5 | EQ6 | EQ7 | EQ8 | EQ9 | EQ5 | EQ6 | EQ7 | EQ8 | EQ9 |
| Strongly disagree | 59% | 9% | 8% | 9% | 24% | 63% | 4% | 9% | 4% | 12% |
| Disagree | 31% | 21% | 22% | 24% | 44% | 29% | 13% | 21% | 19% | 36% |
| Neutral | 7% | 36% | 19% | 42% | 26% | 3% | 23% | 24% | 41% | 37% |
| Agree | 2% | 31% | 41% | 25% | 6% | 4% | 56% | 37% | 34% | 14% |
| Strongly Agree | 0% | 3% | 10% | 1% | 0% | 0% | 4% | 9% | 2% | 1% |

EQ5: *VSAQs are easier than MCQs*.
EQ6: *VSAQs are more in line with daily clinical practice than MCQs.*
EQ7: *I prepare differently for an assessment with VSAQs than for an assessment with MCQs.*
EQ8: *VSAQs would be a better preparation for clinical practice than MCQs*.
EQ9: *Through the use of VSAQs, the test is better aligned with this course, than a test using MCQs.*

**Supplemental table 6.** Median (IQR) scores of the 5-point Likert scale evaluation questions in the formative exam. EQ1-4 halfway of the exam after the MCQs (MCQ*first*) or VSAQs (VSAQ*first*); EQ5-EQ9 at the end of the exam after both MCQs and VSAQs.

| | **Regulation and Metabolism** | | **Diseases of the Abdomen** | |
|---|---|---|---|---|
| | **MCQ*first* (*n* = 104)** | **VSAQ*first* (*n* = 112)** | **MCQ*first* (*n* = 85)** | **VSAQ*first* (*n* = 64)** |
| EQ1 | 4 (3-4) | 4 (3-4) | 4 (3-4) | 4 (4-4) |
| EQ2 | 2 (2-3) | 2 (2-3) | 3 (2-3) | 2 (2-2) |
| EQ3 | 4 (4-4) | 4 (4-5) | 4 (3-4) | 4 (4-5) |
| EQ4 | 6 (5-6) | 6 (6-7) | 5 (4-6) | 6 (5-6) |
| EQ5 | 1 (1-2) | | 1 (1-2) | |
| EQ6 | 3 (2-4) | | 4 (3-4) | |
| EQ7 | 4 (2-4) | | 3 (2-4) | |
| EQ8 | 3 (2-4) | | 3 (2-4) | |
| EQ9 | 2 (2-3) | | 2 (2-3) | |

EQ1: *The questions are a good representation of how I would be expected to answer questions in clinical practice.*
EQ2: *I found the questions easy.*
EQ3: *I was often unsure whether my answer would be correct.*
EQ4: *If I had to give an estimate of the grade I would have achieved based on these questions, my estimate would be <grade>.*
EQ5: *VSAQs are easier than MCQs*.
EQ6: *VSAQs are more in line with daily clinical practice than MCQs.*
EQ7: *I prepare differently for an assessment with VSAQs than for an assessment with MCQs.*
EQ8: *VSAQs would be a better preparation for clinical practice than MCQs*.
EQ9: *Through the use of VSAQs, the test is better aligned with this course, than a test using MCQs.*

**Supplemental table 7**. Mean scores of the 5-point Likert scale evaluation questions after the summative exam.

| | Regulation and Metabolism | | Diseases of the Abdomen | |
|---|---|---|---|---|
| | **Q1 (*n* = 147)** | **Q2 (*n* = 148)** | **Q1 (*n* = 85)** | **Q2 (*n* = 85)** |
| Strongly disagree | 13% | 53% | 26% | 18% |
| Disagree | 26% | 30% | 27% | 33% |
| Neutral | 17% | 12% | 6% | 19% |
| Agree | 35% | 5% | 38% | 26% |
| Strongly Agree | 10% | 0% | 4% | 5% |
| Median (IQR) | 2 (3-4) | 1 (1-2) | 1 (2-4) | 2 (2-4) |

Q1*: Because I knew that I would be tested by VSAQs, I studied in another way than I normally would;*
Q2: *Through the use of VSAQs, the test was a better representation of what I learned in this course, than a test using MCQs.*

**Supplemental table 8**. Mean scores of the 5-point Likert scale questions on constructive alignment after the summative exam (1: strongly disagree, 2: disagree, 3: neutral, 4: agree, 5: strongly agree).

| | Regulation and Metabolism | | | | Diseases of the Abdomen | | | |
|---|---|---|---|---|---|---|---|---|
| | Q1 | | Q2 | | Q1 | | Q2 | |
| | *N* | Mean (SD) | *N* | Mean (SD) | *N* | Mean (SD) | *N* | Mean (SD) |
| '16/'17 | *NA* | *NA* | 197 | 3.6 (0.9) | *NA* | *NA* | 170 | 3.6 (0.9) |
| '17/'18 | 50 | 3.3 (1.1) | 50 | 3.2 (1.1) | 66 | 3.6 (1.0) | 66 | 3.5 (0.7) |
| '18/'19 | 63 | 3.1 (1.2) | 62 | 2.9 (1.2) | 62 | 2.2 (1.1) | 62 | 2.2 (1.1) |
| '20/'21 | 149 | 2.2 (1.1) | 149 | 2.2 (1.1) | 127 | 2.9 (1.1) | 127 | 3.1 (1.1) |

Q1: *The assessment as a whole (form and content) is appropriate for what you should have mastered at the end of the course.*
Q2 = *The (online) test formats matched what I have learned; NA = not available.*
